# Image- vs. histogram-based considerations in semantic segmentation of pulmonary hyperpolarized gas images

**DOI:** 10.1101/2021.03.04.21252588

**Authors:** Nicholas J. Tustison, Talissa A. Altes, Kun Qing, Mu He, G. Wilson Miller, Brian B. Avants, Yun M. Shim, James C. Gee, John P. Mugler, Jaime F. Mata

## Abstract

Magnetic resonance imaging (MRI) using hyperpolarized gases has made possible the novel visualization of airspaces in the human lung, which has advanced research into the growth, development, and pathologies of the pulmonary system. In conjunction with the innovations associated with image acquisition, multiple image analysis strategies have been proposed and refined for the quantification of such lung imaging with much research effort devoted to semantic segmentation, or voxelwise classification, into clinically oriented categories based on ventilation levels. Given the functional nature of these images and the consequent sophistication of the segmentation task, many of these algorithmic approaches reduce the complex spatial image information to intensity-only considerations, which can be contextualized in terms of the intensity histogram. Although facilitating computational processing, this simplifying transformation results in the loss of important spatial cues for identifying salient image features, such as ventilation defects (a well-studied correlate of lung pathophysiology), as spatial objects. In this work, we discuss the interrelatedness of the most common approaches for histogram-based optimization of hyperpolarized gas lung imaging segmentation and demonstrate how certain assumptions lead to suboptimal performance, particularly in terms of measurement precision. In contrast, we illustrate how a convolutional neural network is optimized (i.e., trained) directly within the image domain to leverage spatial information. This image-based optimization mitigates the problematic issues associated with histogram-based approaches and suggests a preferred future research direction. Importantly, we provide the entire processing and evaluation framework, including the newly reported deep learning functionality, as open-source through the well-known Advanced Normalization Tools ecosystem.

## 1 Introduction

### 1.1 Historical overview of quantification

Early attempts at quantification of ventilation images were limited to enumerating the number of ventilation defects or estimating the proportion of ventilated lung (35, 61, 62) which has evolved to more sophisticated techniques used currently. A brief outline of major contributions can be roughly sketched to include:

- binary thresholding based on relative intensities (42, 55),
- linear intensity standardization based on a global rescaling of the intensity histogram to a reference distribution based on healthy controls, i.e., “linear binning” (53, 54),
- nonlinear intensity standardization based on piecewise affine transformation of the intensity histogram using a customized hierarchical (34, 52) or adaptive (8) k-means algorithm,
- nonlinear intensity standardization using fuzzy c-means (4) with spatial considerations based on local voxel neighborhoods (7), and
- Gaussian mixture modeling (GMM) of the intensity histogram with Markov random field (MRF) spatial prior modeling (64).

An early semi-automated technique used to compare smokers and never-smokers relied on manually drawn regions to determine a threshold based on the mean signal and noise values (55). Related approaches, which use a simple rescaled threshold value to binarize the ventilation image into ventilated and non-ventilated regions (33), continue to find modern application (42). Similar to the histogram-only algorithms (i.e., linear binning and hierarchical k-means, discussed below), these approaches do not take into account the various MRI artefacts such as noise (43, 44) and the intensity inhomogeneity field (60) which prevent hard threshold values from distinguishing tissue types precisely consistent with that of human experts. In addition, to provide a more granular categorization of ventilation for greater compatibility with clinical qualitative assessment, many current techniques have increased the number of voxel classes (i.e., clusters) beyond the binary categories of “ventilated” and “non-ventilated.”

Linear binning is a simplified type of MR intensity standardization (51) in which images from healthy controls are normalized to the range [0, 1] and then used to calculate the cluster intensity boundary values, based on an aggregated estimate of the parameters of a single Gaussian fit. Subject images to be segmented are then rescaled to this reference histogram (i.e., a global affine 1-D transform). This mapping results in alignment of the cluster boundaries such that corresponding labels are assumed to have similar clinical interpretation. In addition to the previously mentioned limitations associated with hard threshold values, such a global transform does not account for MR intensity nonlinearities that have been well-studied (47–51) and are known to cause significant intensity variation even in the same region of the same subject. As stated in (47):

> Intensities of MR images can vary, even in the same protocol and the same sample and using the same scanner. Indeed, they may depend on the acquisition conditions such as room temperature and hygrometry, calibration adjustment, slice location, B0 intensity, and the receiver gain value. The consequences of intensity variation are greater when different scanners are used.

As we illustrate in subsequent sections, ignoring these nonlinearities is known to have significant consequences in the well-studied (and somewhat analogous) area of brain tissue segmentation in T1-weighted MRI (e.g., (45, 46, 59)). Here we demonstrate its effects in hyperpolarized gas imaging quantification robustness in conjunction with noise considerations. In addition, the reference distribution required by linear binning assumes sufficient agreement as to what constitutes a “healthy control,” whether a Gaussian fit is appropriate, and, even assuming the latter, whether or not the parameter values can be combined in a linear fashion to constitute a single reference standard. Of additional concern, though, is that the requirement for a healthy cohort for determination of algorithmic parameters introduces a non-negligible source of measurement variance, as we will also demonstrate.

Previous attempts at histogram standardization (50, 51) in light of MR intensity nonlinearities have relied on 1-D piecewise affine mappings between corresponding structural features found within the histograms themselves (e.g., peaks and valleys). For example, structural MRI, such as T1-weighted neuroimaging, utilizes the well-known relative intensities of major tissue types (i.e., cerebrospinal fluid (CSF), gray matter (GM), and white matter (WM)), which characteristically correspond to visible histogram peaks, as landmarks to determine the nonlinear intensity mapping between histograms. However, in hyperpolarized gas imaging of the lung, no such characteristic structural features exist, generally speaking, between histograms. Additionally, because of the functional nature of these images, the segmentation clusters that correspond to features of interest are not necessarily guaranteed to exist (e.g., ventilation defects in the case of healthy normal subjects with no lung pathology).

The approach used by some groups (26, 52) of employing some variant of the well-known k-means algorithm as a clustering strategy (41) to minimize the within-class variance of its intensities can be viewed as an alternative optimization strategy for determining a nonlinear mapping between histograms for a type of MR intensity standardization. K-means constitutes an algorithmic approach with additional flexibility and sophistication over linear binning as it employs prior knowledge in the form of a generic clustering desideratum for optimizing a type of MR intensity standardization.^1^

Similar to k-means, fuzzy c-means seeks to minimize the within-class sample variance but includes a per-sample membership weighting (6). Later innovations included the incorporation of spatial considerations using class membership values of the local voxel neighborhood (5). Both k-means and fuzzy spatial c-means were compared for segmentation of hyperpolarized 3He and 129Xe images in (7) with the latter evidencing improved performance over the former which is due, at least in part, to the additional spatial considerations. Despite relatively good performance, however, fuzzy c-means also seeks cluster membership in the histogram (i.e., intensity-only) domain with only simplistic neighborhood modeling during optimization.

Histogram-based optimization is used in conjunction with spatial considerations in the segmentation algorithm detailed in (64). This algorithm is based on a well-established iterative approach originally used for NASA satellite image processing and subsequently appropriated for brain tissue segmentation in (40). A Gaussian mixture model (GMM) is used to model the intensity clusters of the histogram with class modulation in the form of probabilistic voxelwise label considerations, i.e., Markov random field (MRF) modeling, within image neighborhoods (32) optimized with the expectation-maximization (EM) algorithm (31). This has the advantage, in contrast to histogram-only algorithms, that it softens the intensity thresholds between class labels which demonstrates robustness to certain imaging distortions, such as noise. However, as we will demonstrate, this algorithm is also flawed in the inherent assumption that meaningful structure is found, and can be adequately characterized, within the associated image histogram in order to optimize a multi-class labeling. In particular, this algorithm is susceptible to MR nonlinear intensity artefacts.

Additionally, many of these segmentation algorithms use N4 bias correction (63), an extension of the nonuniform intensity normalization (N3) algorithm (60), to mitigate MR intensity inhomogeneity artefacts. Interestingly, N3/N4 also iteratively optimizes towards a final solution using information from both the histogram and image domains. Based on the intuition that the bias field acts as a smoothing convolution operation on the original image intensity histogram, N3/N4 optimizes a nonlinear (i.e., deformable) intensity mapping, based on histogram deconvolution. This nonlinear mapping is constrained such that its effects smoothly vary across the image. Additionally, due to the deconvolution operation, this nonlinear mapping sharpens the histogram peaks which presumably correspond to tissue types. While such assumptions are appropriate for the domain in which N3/N4 was developed (i.e., T1-weighted brain tissue segmentation) and while it is assumed that the enforcement of low-frequency modulation of the intensity mapping prevents new image features from being generated, it is not clear what effects N4 parameter choices have on the final segmentation solution, particularly for those algorithms that are limited to intensity-only considerations and not robust to the aforementioned MR intensity nonlinearities.

### 1.2 Motivation for current study

Investigating the assumptions outlined above, particularly those associated with the nonlinear intensity mappings due to both the MR acquisition and inhomogeneity mitigation preprocessing, we became concerned by the susceptibility of the histogram structure to such variations and the potential effects on current clinical measures of interest derived from these algorithms (e.g., ventilation defect percentage). Specifically, we developed the ability to simulate such MR nonlinear intensity variation by warping the intensity histogram and propagating the intensity changes to the original image. Details as to the availability and code functionality is provided below in the Methods section. Suffice it to say, we noticed that histogram-based intensity perturbations can produce virtually little, if any, changes in the features of the image despite a relatively significant change in the histogram structure. Such effects imply that MR artefacts could profoundly impact histogram-based algorithmic performance. Figure 1 provides a sample visualization representing some of the structural changes that we observed when simulating these nonlinear mappings. It is important to notice that even relatively small alterations in the image intensities can have significant effects on the histogram even though a visual assessment of the image can remain largely unchanged.

**Figure 1:**
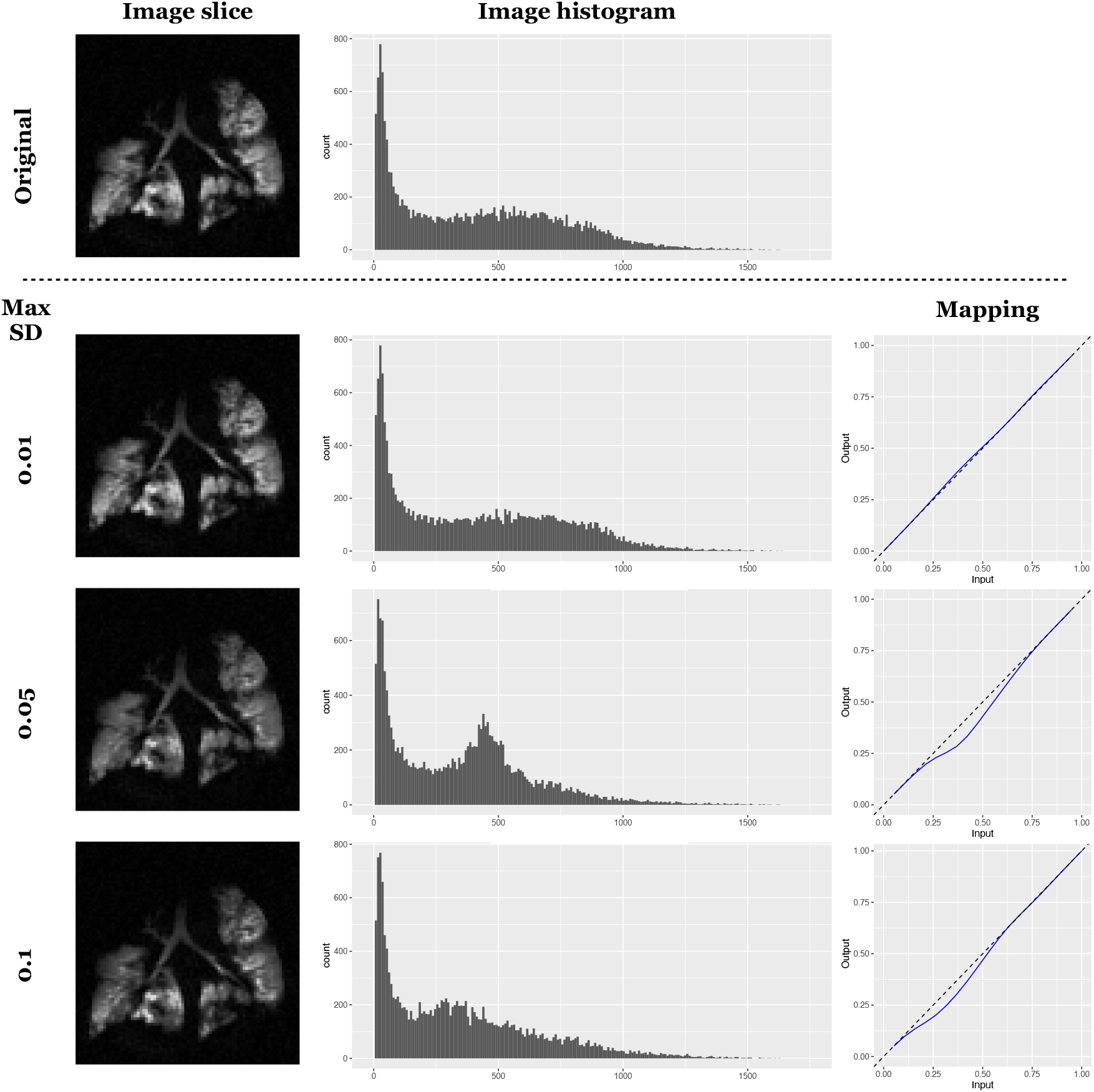
Illustration of the effect of MR nonlinear intensity warping on the histogram structure where the amount of random 1-D deformation increases with each row. By simulating these types of intensity changes, we can visualize the effects on the underlying intensity histograms and investigate the effects on salient outcome measures. Here we simulate intensity mappings which, although relatively small, can have a significant effect on the histogram structure.

To briefly explore these effects further for the purposes of motivating additional experimentation, we provide a summary illustration from a set of image simulations in Figure 2 which are detailed later in this work and used for algorithmic comparison. Simulated MR artefacts were applied to each image which included both noise and nonlinear intensity mappings (and their combination) using two separate data sets: one in-house data set consisting of 51 129Xe gas lung images and the publicly available data described in (3) and made available at Harvard’s Dataverse online repository (2) consisting of 29 hyperpolarized gas lung images. These two data sets resulted in a total simulated cohort of 51 + 29 = 80 images (*×*10 simulations per image *×*3 types of artefact simulations). Prior to any algorithmic comparative analysis, we quantified the difference of each simulated image with the corresponding original image using the structural similarity index measurement (SSIM) (25). SSIM is a highly cited measure which quantifies structural differences between a reference and distorted (i.e., transformed) image based on known properties of the human visual system. SSIM has a range [*−*1, 1] where 0 indicates no structural similarity and 1 indicates perfect structural similarity. We also generated the histograms corresponding to these images. Although several histogram similarity measures exist, we chose Pearson’s correlation primarily as it resides in the same min/max range as SSIM with analogous significance. In addition to the fact that the image-to-histogram transformation discards important spatial information, from Figure 2 it should be apparent that this transformation also results in greater variance in the resulting information under common MR imaging artefacts, according to these measures. Thus, prior to any algorithmic considerations, these observations strongly suggest that optimizing in the domain of the histogram will be generally less informative and less robust than optimizing directly in the image domain.

**Figure 2:**
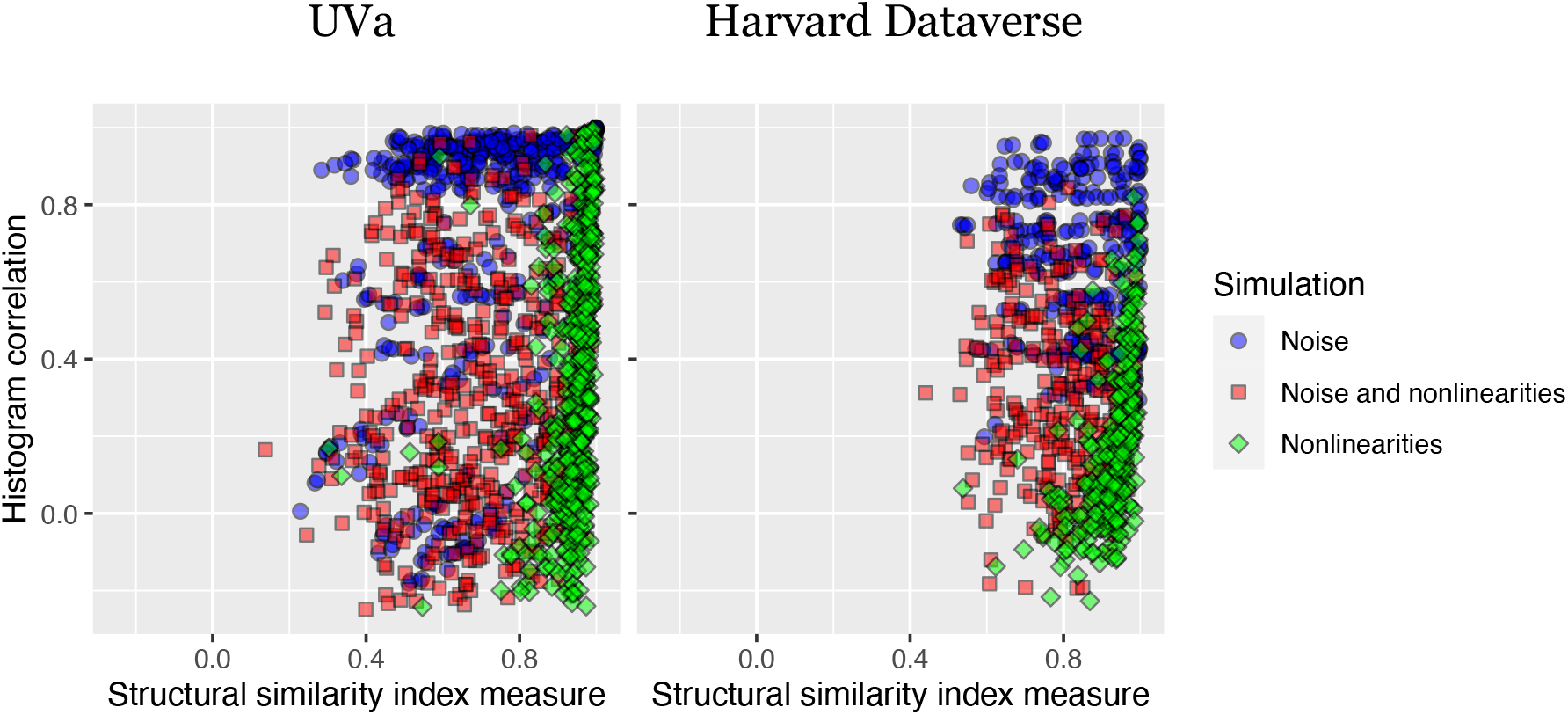
Multi-site: (left) University of Virginia (UVa) and (right) Harvard Dataverse 129Xe data. Image-based SSIM vs. histogram-based Pearson’s correlation differences under distortions induced by the common MR artefacts of noise and intensity nonlinearities. For the nonlinearity-only simulations, the images maintain their structural integrity as the SSIM values remain close to 1. This is in contrast to the corresponding range in histogram similarity which is much larger. The effects with simulated Gaussian noise are similar where the range in histogram differences with simulated noise is much greater than the range in SSIM. Both sets of observations are evidence of the lack of robustness to distortions in the histogram domain in comparison with the original image domain.

Ultimately, we are not claiming that these algorithms are erroneous, per se. Much of the relevant research has been limited to quantifying differences with respect to ventilation versus non-ventilation in various clinical categories and these algorithms have certainly demonstrated the capacity for advancing such research. Furthermore, as the sample segmentations in Figure 3 illustrate, when considered qualitatively, each segmentation algorithm appears to produce a reasonable segmentation even though the voxelwise differences are significant (as are the corresponding histograms). However, the aforementioned artefact issues influence quantitation in terms of core scientific measurement principles such as precision (e.g., reproducibility and repeatability (8, 30)) and bias which are obscured in isolated considerations but become increasingly significant with multi-site (24) and large-scale studies. In addition, generally speaking, refinements in measuring capabilities correlate with scientific advancement so as acquisition and analysis methodologies improve, so should the level of sophistication and performance of the underlying measurement tools.

**Figure 3:**
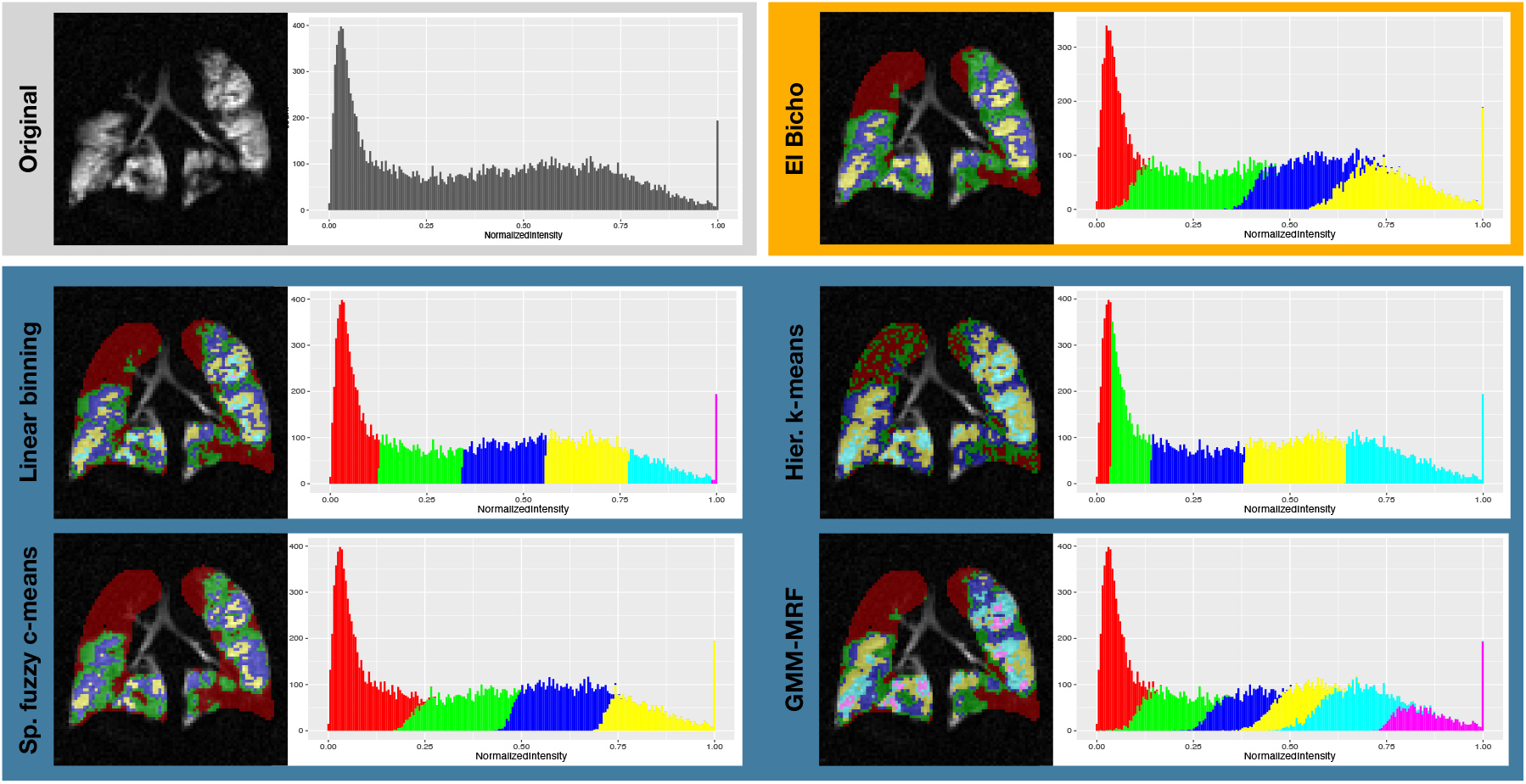
Illustration of sample segmentations produced by the four algorithms described above (i.e., linear binning, hierarchical k-means, spatial fuzzy c-means, and GMM-MRF) and the deep learning algorithm (“El Bicho”) described below on a single cystic fibrosis subject. Also included are the corresponding segmentation histograms. Although quite disparate in the actual labeling of the lung and resulting histogram, each algorithm produces a reasonable parcellation.

In assessing these segmentation algorithms for hyperpolarized gas imaging, it is important to note that human expertise leverages more than relative intensity values to identify salient, clinically relevant features in images—something more akin to the complex structure of deep-layered neural networks (29), particularly convolutional neural networks (CNN). Such models have demonstrated outstanding performance in certain computational tasks, including classification and semantic segmentation in medical imaging (28). Their potential for leveraging spatial information from images surpasses the perceptual capabilities of previous approaches and even rivals that of human raters (58). Importantly, CNN optimization occurs directly in the image space to learn complex spatial features, in contrast to the previously discussed methods where optimization (primarily) concerns image intensity only information. We introduced a deep learning approach in (56) and further expand on that work for comparison with existing approaches below. Although we find its performance to be quite promising, more fundamental to this work than the network itself is simply pointing to the general potential associated with deep learning for analyzing hyperpolarized gas images *as spatial samplings of real-world objects*, as opposed to lossy representations of such objects. In the spirit of open science, we have made the entire evaluation framework, including our novel contributions, available within the Advanced Normalization Tools software ecosystem (ANTsX) (39).

## 2 Materials and methods

### 2.1 Hyperpolarized gas imaging acquisition

#### 2.1.1 University of Virginia cohort

A retrospective dataset was collected consisting of young healthy (*n* = 10), older healthy (*n* = 7), cystic fibrosis (CF) (*n* = 14), interstitial lung disease (ILD) (*n* = 10), and chronic obstructive pulmonary disease (*n* = 10). MR imaging with hyperpolarized 129Xe gas was performed under an Institutional Review Board (IRB) approved protocol with written informed consent obtained from each subject. In addition, all imaging was performed under a Food and Drug Administration (FDA) approved physician’s Investigational New Drug application. MRI data were acquired on a 1.5 T whole-body MRI scanner (Siemens Avanto, Siemens Medical Solutions, Malvern, PA) with broadband capabilities and a flexible 129Xe chest radiofrequency coil (RF; IGC Medical Advances, Milwaukee, WI; or Clinical MR Solutions, Brookfield, WI). During a *≤* 10 breath-hold following the inhalation of *≈* 1000 mL of hyperpolarized 129Xe mixed with nitrogen up to a volume equal to 1/3 forced vital capacity (FVC) of the respective subject, a set of 15-17 contiguous coronal lung slices were collected in order to cover the entire lungs. Parameters of the gradient echo (GRE) sequence with a spiral k-space sampling with 12 interleaves for 129Xe MRI were as follows: repetition time msec / echo time msec, 7/1; flip angle, 20°; matrix, 128 *×* 128: in-plane voxel size, 4 *×* 4 mm; section slice thickness, 15 mm; and intersection gap, none. The data were deidentified prior to analysis.

#### 2.1.2 Harvard Dataverse cohort

In addition to these data acquired at the University of Virginia, we also processed a publicly available lung dataset (2) available at the Harvard Dataverse and detailed in (3). These data comprised the original 129Xe acquisitions from 29 subjects (10 healthy controls and 19 mild intermittent asthmatic individuals) with corresponding lung masks. In addition, seven artificially SNR-degraded images per acquisition were also part of this data set but not used for the analyses reported below. The image headers were corrected for proper canonical anatomical orientation according to Nifti standards and uploaded to the GitHub repository associated with this work.

### 2.2 Algorithmic implementations

In support of the discussion in the Introduction, we performed various experiments to compare the algorithms described previously, viz. linear binning (54), hierarchical k-means (52), fuzzy spatial c-means (7), GMM-MRF (specifically, ANTs-based Atropos tailored for functional lung imaging) (64), and a trained CNN with roots in our earlier work (56), which we have dubbed “El Bicho.”^2^ A fair and accurate comparison between algorithms necessitates several considerations which have been outlined previously (65). In designing the evaluation study:

- All algorithms and evaluation scripts have been implemented using open-source tools by the first author. The linear binning and hierarchical k-means algorithms were recreated using existing R functionality. These have been made available as part of the GitHub repository corresponding to this work.^3^ Similarly, N4, fuzzy spatial c-means, Atropos-based lung segmentation, and the trained CNN approach are all available through ANTsR/ANTsRNet: `ANTsR::n4BiasFieldCorrection, ANTsR::fuzzySpatialCMeansSegmentation, ANTsR::functionalLungSegmentation`, and `ANTsRNet::elBicho`, respectively. Python versions are also available through ANTsPy/ANTsPyNet. The trained weights for the CNN are publicly available and are automatically downloaded when running the program.
- The University of Virginia imaging data used for the evaluation is available upon request and through a data sharing agreement. In addition to the citation providing the online location of the original Harvard Dataverse data, a header-modified version of these data is available in the GitHub repository associated with this manuscript. Additional evaluation plots have also been made available.
- An extremely important algorithmic hyperparameter is the number of ventilation clusters. In order to minimize differences in our set of evaluations, we merged the number of resulting clusters, post-optimization, to only three clusters: “ventilation defect,” “hypo-ventilation,” and “other ventilation” where the first two clusters for each output are the same as the original implementations and the remaining clusters are merged into the third category (i.e., “other ventilation”). It is important to note that none of the evaluations use these categorical definitions in a cross-algorithmic fashion. They are only used to assess within-algorithm consistency.
- A significant issue was whether or not to use the N4 bias correction algorithm as a preprocessing step. We ultimately decided to include it for two reasons.^4^ First, it is explicitly used in multiple algorithms (e.g., (8, 18, 42, 54, 64)) despite the issues raised previously due to the fact that it qualitatively improves image appearance.^5^ Another practical consideration for N4 preprocessing was due to the parameters of the reference distribution required by the linear binning algorithm. Additional details are provided in the Results section.

### 2.3 Introduction of the image-based “El Bicho” network

We extended the deep learning functionality first described in (56) to improve performance and provide a more clinically granular labeling (i.e., four clusters here instead of two in the previous work). In addition, further modifications incorporated additional data during training, added attention gating (36) to the U-net network (57) along with recommended hyperparameters (38), and a novel data augmentation strategy.

#### 2.3.1 Network training

“El Bicho” is a 2-D U-net network which was trained with several parameters recommended by recent exploratory work (38). The images are sufficiently small such that 3-D training is possible. However, given the large voxel anisotropy for much of our data (both coronal and axial), we found a 2-D approach to be sufficient. Nevertheless, a 2.5-D approach is an optional way to run the code for isotropic data where network prediction can occur in more than one slice direction and the results subsequently averaged. Four total network layers were employed with 32 filters at the base layer which was doubled at each subsequent layer. Multiple training runs were performed where initial runs employed categorical cross entropy as the loss function. Upon convergence, training continued with the multi-label Dice function (37)

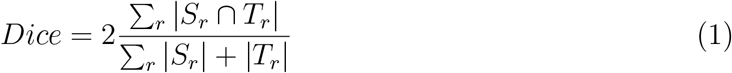

where *S*_*r*_ and *T*_*r*_ refer to the source and target regions, respectively.

Training data (using an 80/20—training/testing split) was composed of the ventilation image, lung mask, and corresponding ventilation-based parcellation. The lung parcellation comprised four labels based on the Atropos ventilation-based segmentation (64). Six clusters were used to create the training data and combined to four for training the CNN. In using this GMM-MRF algorithm (which is the only one to use spatial information in the form of the MRF prior), we attempt to bootstrap a superior network-based segmentation approach by using the encoder-decoder structure of the U-net architecture as a dimensionality reduction technique. None of the evaluation data used in this work were used as training data. Responses from two subjects at the last layer of the network (with *n* = 32 filters) are illustrated in Figure 5.

A total of five random slices per image were selected in the acquisition direction (both axial and coronal) for inclusion within a given batch (batch size = 128 slices). Prior to slice extraction, both random noise and randomly-generated, nonlinear intensity warping was added to the 3-D image (see Figure 4) using ANTsR/ANTsRNet functions (`ANTsR::addNoiseToImage,` and `ANTsRNet::histogramWarpImageIntensities`) with analogs in ANTsPy/ANTsPyNet. 3-D images were intensity normalized to have 0 mean and unit standard deviation. The noise model was additive Gaussian with 0 mean and a randomly chosen standard deviation value between [0, 0.3]. Histogram-based intensity warping used the default parameters. These data augmentation parameters were chosen to provide realistic but potentially difficult cases for training. In terms of hardware, all training was done on a DGX (GPUs: 4X Tesla V100, system memory: 256 GB LRDIMM DDR4).

**Figure 4:**
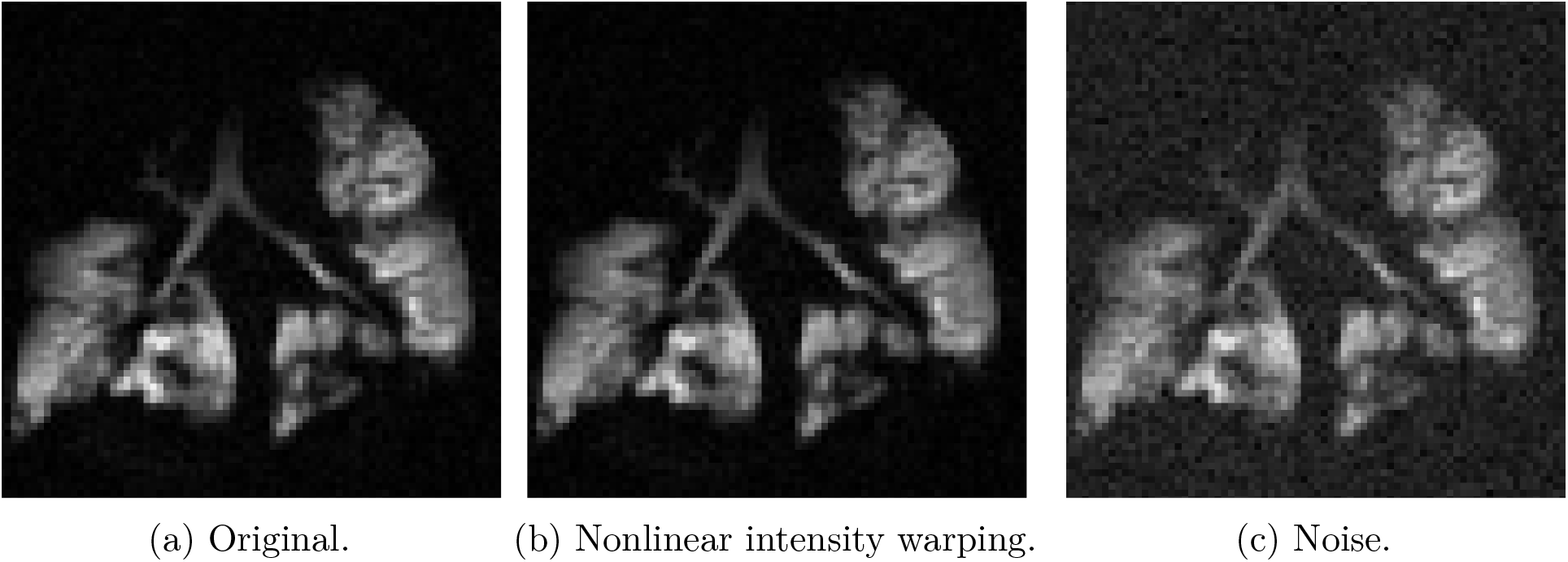
Custom data augmentation strategies for training to force a solution which focuses on the underlying ventilation-based lung structure. (b) Nonlinear intensity warping based on smoothly varying perturbations of the image histogram. (c) Additive Gaussian noise included for increasing the robustness of the segmentation network.

**Figure 5:**
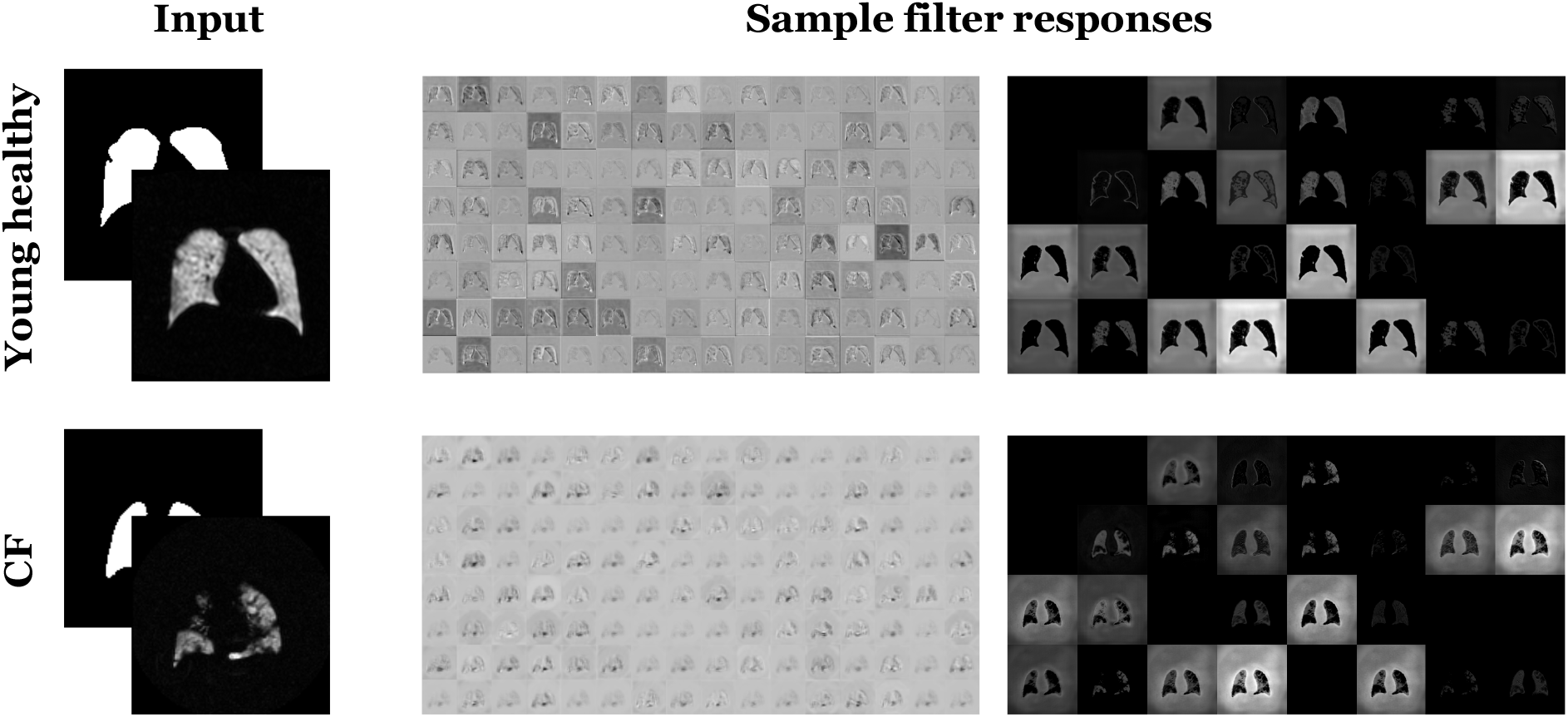
Optimized feature responses from both the encoding and decoding branches of the U-net network generated from a (top) young healthy subject and (bottom) CF patient. Note that these are optimized responses which take advantage of both the intensities and their spatial relationships.

#### 2.3.2 Pipeline processing

An example R-based code snippet is provided in Listing 1 demonstrating how to process a single ventilation image using `ANTsRNet::elBicho`. If a simultaneous proton image has been acquired, `ANTsRNet::lungExtraction` can be used to generate the requisite lung mask input. As mentioned previously, by default the prediction occurs slice-by-slice along the direction of anisotropy. Alternatively, prediction can be performed in all three canonical directions and averaged to produce the final solution.

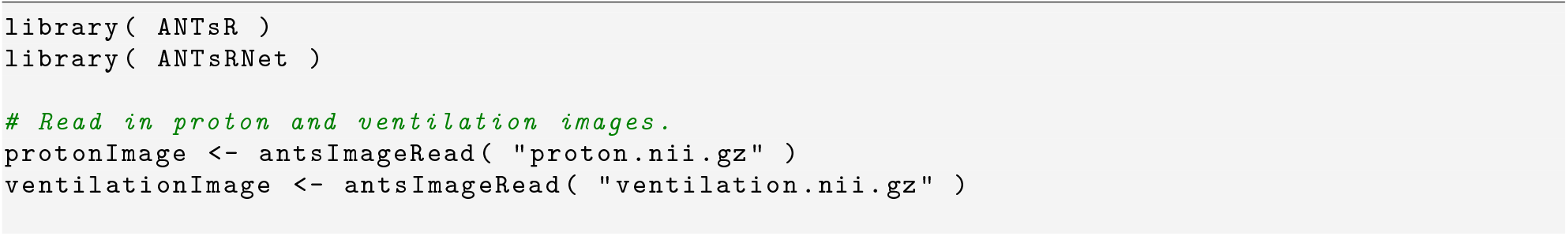

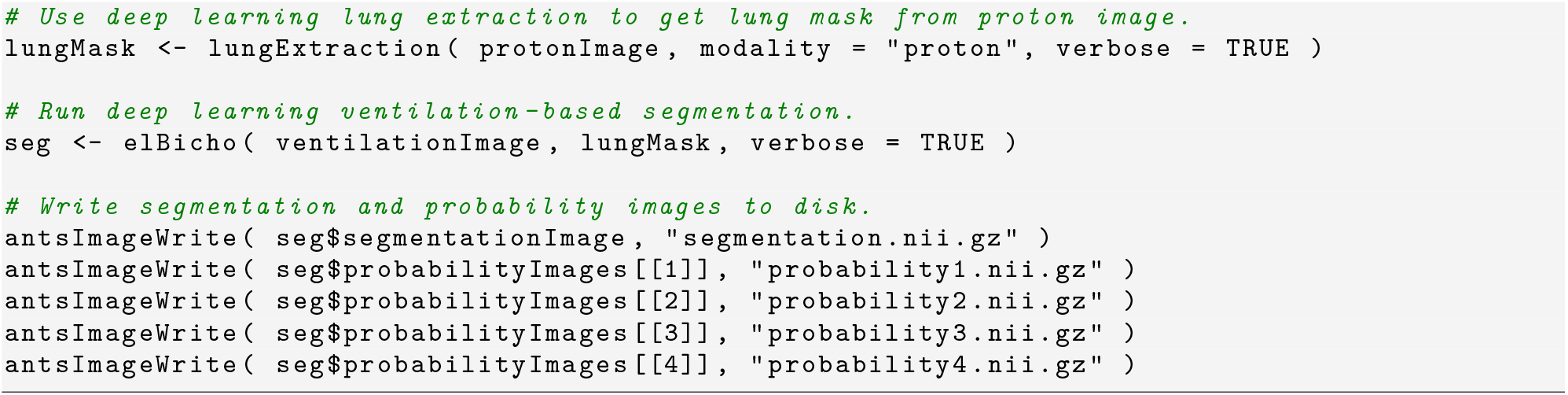

Listing 1: ANTsR/ANTsRNet command calls for processing a single ventilation image using El Bicho.

## 3. Results

We performed several comparative evaluations to probe the previously mentioned algorithmic issues which are broadly categorized in terms of measurement bias and precision, with most of the focus being on the latter. Given the lack of ground-truth in the form of segmentation images, addressing issues of measurement bias is difficult. In addition to the fact that the number of ventilation clusters is not consistent across algorithms, it is not clear that the ventilation categories across algorithms have identical clinical definition. This prevents application of various frameworks accommodating the lack of ground-truth for segmentation performance analysis (e.g., (27)) to these data.

As we mentioned in the Introduction, all the algorithms have demonstrated research utility and potential clinical utility based on findings using derived measures. This is supported by our first evaluation which is based on diagnostic prediction of given clinical categories assigned to the imaging cohort using derived random forest models (21). This approach also provides an additional check on the validity of the algorithmic implementations. However, it is important to recognize that this evaluation is extremely limited as the underlying data are gross measures which do not provide accuracy estimates on the level of the algorithmic output (i.e., voxelwise segmentation).

Having established the general validity of the gross algorithmic output, we then switch to our primary focus which is the comparison of measurement precision between algorithms. We first analyzed the unique requirement of a reference distribution for the linear binning algorithm. The latter is motivated qualitatively through the analogous application of T1-weighted brain MR segmentation. This component is strictly qualitative as the visual evidence and previous developmental history within that field should be sufficiently compelling in motivating subsequent quantitative exploration with hyperpolarized gas lung imaging. These qualitative results segue to quantification of the effects of the choice of reference cohort on the clustering parameters for the linear binning algorithm. We then incorporated the trained El Bicho model in exploring additional aspects of measurement variance based on simulating both MR noise and intensity nonlinearities.

So, in summary, we performed the following evaluations/experiments:^6^

- Global algorithmic bias (in the absence of ground truth)
  – Diagnostic prediction
- Voxelwise algorithmic precision
  – Three-tissue T1-weighted brain MRI segmentation (qualitative analog)
  – Input/output variance based on reference distribution (linear binning only)
  – Effects of simulated MR artefacts on multi-site data

### 3.1 Diagnostic prediction

Due to the absence of ground-truth, we adopted the strategy from previous work (20, 39) where we used cross-validation to build and compare prediction models from data derived from the set of segmentation algorithms. Specifically, we use pathology diagnosis (i.e., “CF,” “COPD,” and “ILD”) as an established research-based correlate of ventilation levels from hyperpolarized gas imaging (e.g., (17–19)) and quantified the predictive capabilities of corresponding binary random forest classifiers (21) of the form:

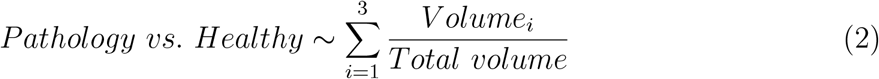

where *Volume*_*i*_ is the volume of the *i*^*th*^ cluster and *Total volume* is total lung volume. We used a training/testing split of 80/20. Due to the small number of subjects, we combined the young and old healthy data into a single category. 100 permutations were used where training/testing data were randomly assigned and the corresponding random forest model was constructed at each permutation.

The resulting receiver operating characteristic (ROC) curves for each algorithm and each diagnostic scenario are provided in Figure 6. In addition, we provide the summary area under the ROC curve (AUC) values in Table 1. In the absence of ground truth, this type of evaluation does provide evidence that all these algorithms produce measurements which are clinically relevant although, it should be noted, that this is a very coarse assessment strategy given the global measures used (i.e., cluster volume percentage) and the general clinical categories employed. In fact, even spirometry measures can be used to achieve highly accurate diagnostic predictions with machine learning techniques (22).

**Table 1:** AUC values describing the algorithmic performance for each set of binary classification simulations: CF vs. Healthy, COPD vs. Healthy, and ILD vs. Healthy. All four algorithms perform significantly better than a random classifier.

|  | CF/Healthy | COPD/Healthy | ILD/Healthy |
| --- | --- | --- | --- |
| **Linear binning** | 0.92 | 0.89 | 0.83 |
| **Hier. k-means** | 0.95 | 0.87 | 0.73 |
| **Spatial fuzzy c-means** | 0.73 | 0.72 | 0.62 |
| **GMM-MRF** | 0.84 | 0.82 | 0.64 |
| **El Bicho** | 0.90 | 0.94 | 0.74 |

**Figure 6:**
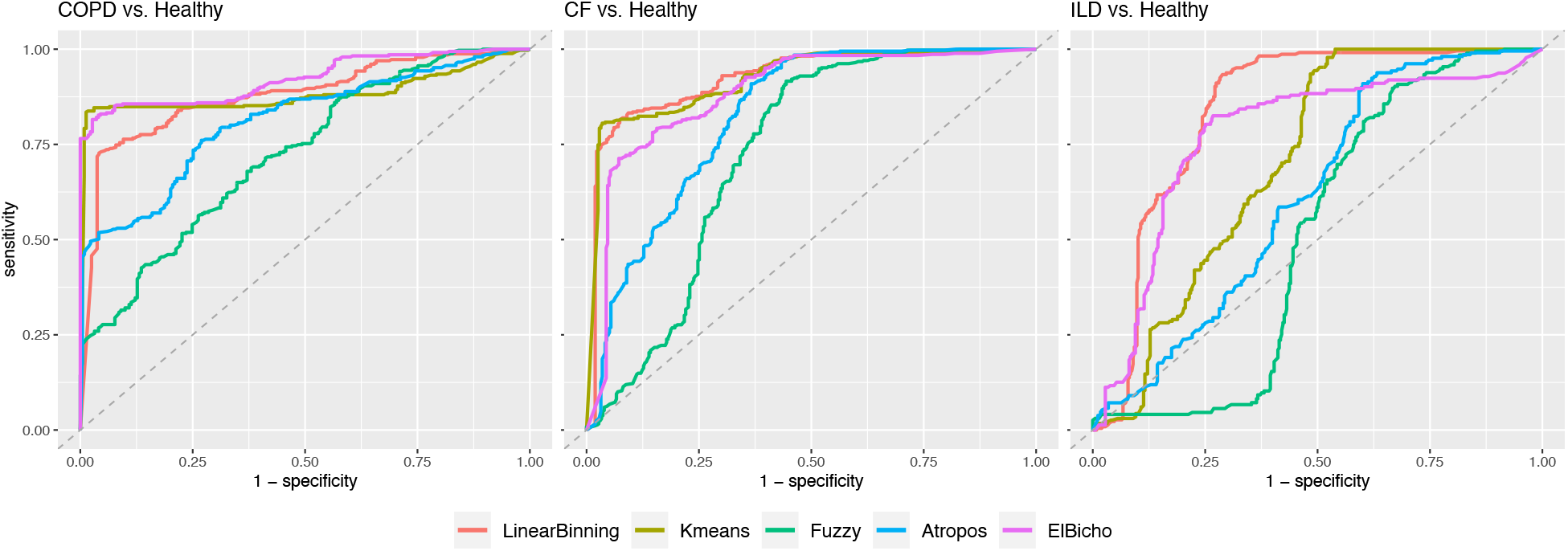
ROC curves resulting from the diagnostic prediction evaluation strategy involving randomly permuted training/testing data sets and predictive random forest models. Summary values are provided in Table 1.

### 3.2 T1-weighted brain segmentation analogy

Much of the quantitative image analysis strategies that have been used for hyperpolarized gas imaging draw on inspiration from fields with a much greater historical background of development, including T1-weighted brain MRI tissue segmentation. The depth of this development can be gauged simply by the number of technical reviews (e.g., (14–16)) and evaluation studies (e.g., (12, 13)) that date back decades. In addition to technical insight, this particular application provides a useful analogy for some of the algorithmic issues discussed and provides context for subsequent evaluations specific to hyperpolarized gas imaging.

In the style of linear binning, we randomly selected ten structurally healthy controls from the publicly available SRPB data set (11) comprising over 1600 participants from 12 sites. After intensity truncation at the 0.99 quantile, we normalize the intensity histogram to [0,1]. Eight of these histograms are provided in the upper left of Figure 7. As we mentioned previously, the histograms for these structural MRI are typically characterized by three peaks which correspond to the CSF, GM, and WM. However, even when normalized to [0, 1] (i.e., global affine mapping), it is obvious that these histogram features do not line up and this is due to the intensity distortion caused by various MR acquisition artefacts mentioned previously. This is an argument from analogy against one of the principal assumptions of linear binning where it is assumed that tissue types (“structural” in the case of T1-weighted brain MRI or “ventilated” in the case of hyperpolarized gas imaging) can be sufficiently aligned with a global rescaling of intensity values. If we pursue this analogy further and use the aggregated reference distribution to segment a different subject, we can see that, in this particular case, whereas the optimization criterion leveraged by k-means and GMM-MRF provide an adequate segmentation, the misalignment in cluster boundaries yield a significant overestimation of the gray matter volume. In the case of hyperpolarized gas images, similar misalignments could cause under-or overestimation of ventilation-based cluster volumes although, in this case, the error is much less obvious given the lack of prior knowledge of functional (vs. anatomical) information.

**Figure 7:**
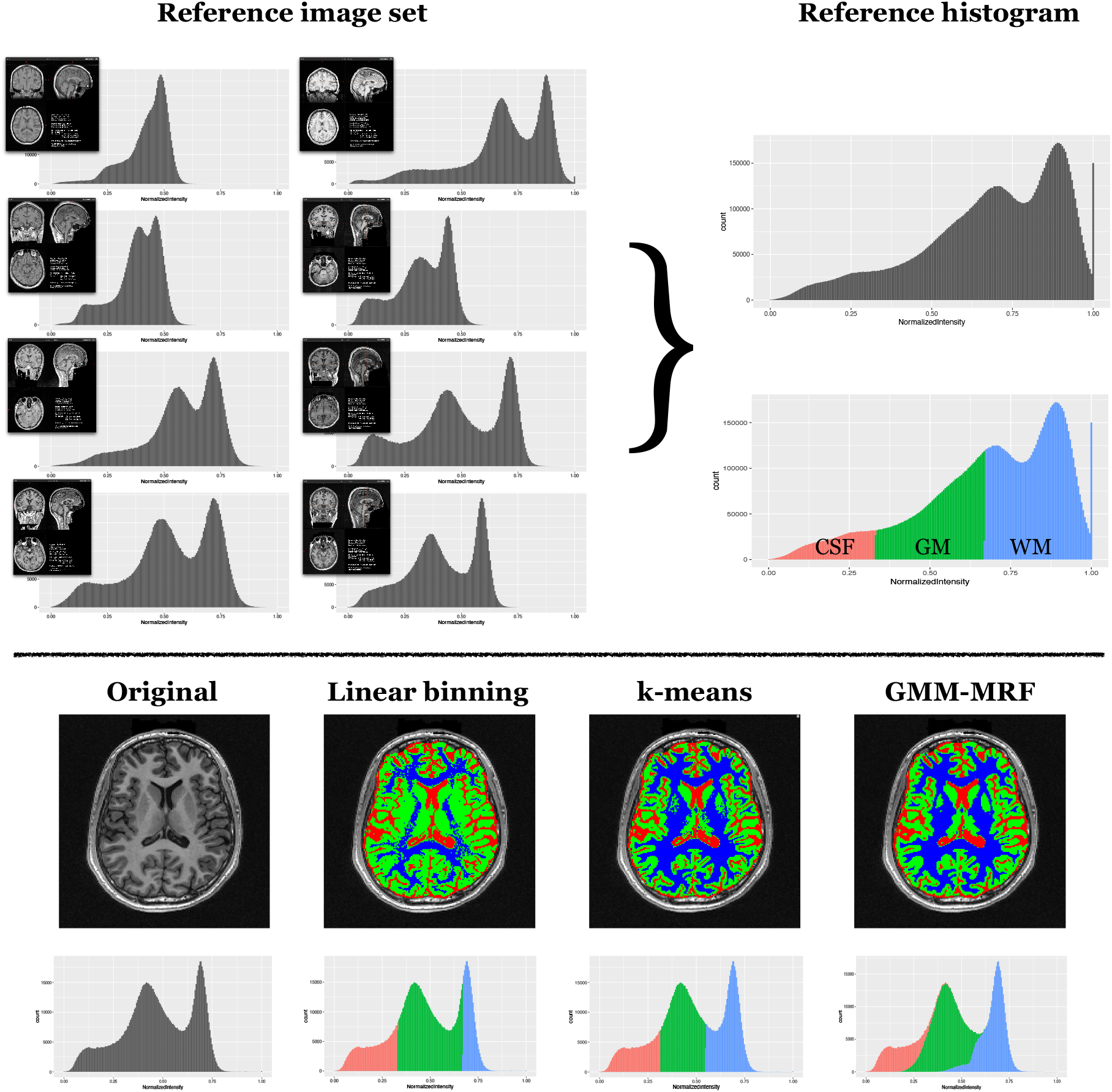
T1-weighted three-tissue brain segmentation analogy. Placing three of the five segmentation algorithms (i.e., linear binning, k-means, and GMM-MRF) in the context of brain tissue segmentation provides an alternative perspective for comparison. In the style of linear binning, we randomly select an image reference set using structurally normal individuals which is then used to create a reference histogram. (Bottom) For a subject to be processed, the resulting hard threshold values yield the linear binning segmentation solution as well as the initialization cluster values for both the k-means and GMM-MRF segmentations which are qualitatively different.

**Figure 8:**
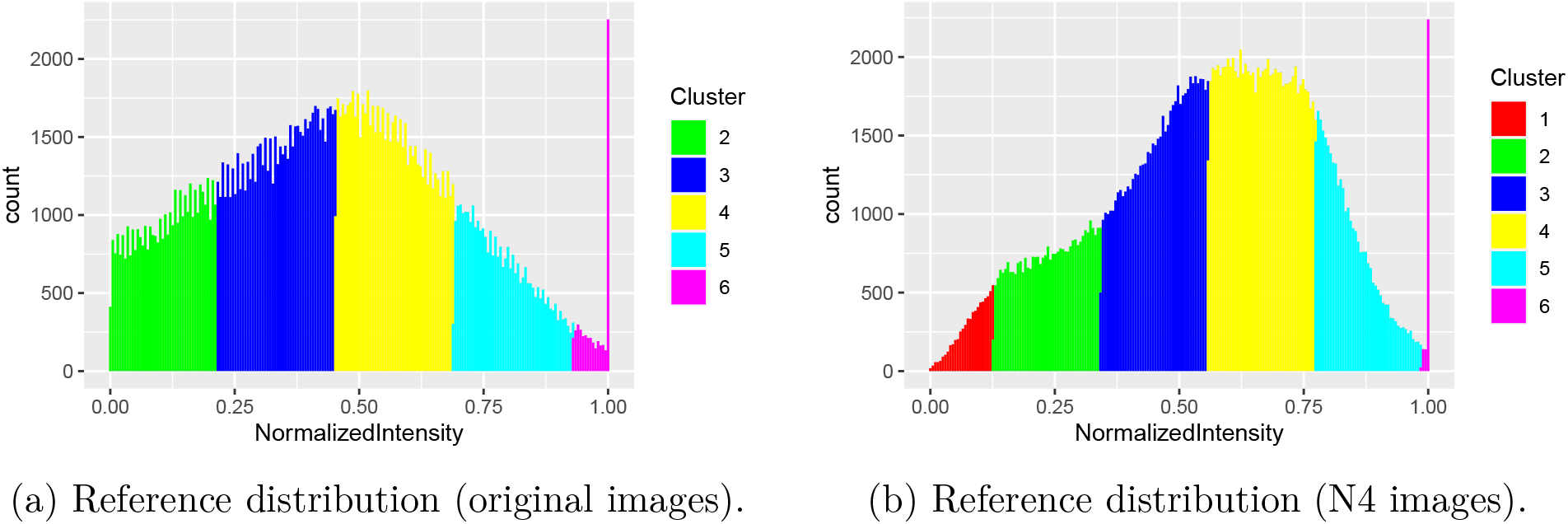
Ten young healthy subjects were combined to create two reference distributions, one based on the (a) original images and the other using (b) N4 preprocessing. Based on the generated mean and standard deviation of the aggregated samples, we label the resulting clusters in the respective histograms. Due to the lower mean and higher standard deviation of the original image set, Cluster 1 is not within the range of [0, 1] for the resulting reference distribution which motivated the use of the N4 preprocessed image set.

### 3.3 Effect of reference image set selection

One of the additional input requirements for linear binning over the other algorithms is the generation of a reference distribution. Therefore we additionally investigated the influence of reference data set on the outcome of linear binning classification, since this is an integral aspect unique to this method. In addition to the output measurement variation caused by choice of the reference image cohort, this played a role in determining whether or not to use N4 preprocessing. As mentioned, a significant portion of N4 processing involves the deconvolution of the image histogram to sharpen the histogram peaks which decreases the standard deviation of the intensity distribution and can also result in a histogram shift. Using the original set of 10 young healthy data with no N4 preprocessing, we created a reference distribution according to (54), which resulted in an approximate distribution of 𝒩 (0.45, 0.24). This produced 0 voxels being classified as belonging to Cluster 1 (Figure 9) because two standard deviations from the mean is less than 0 and Cluster 1 resides in the region below −2 standard deviations. However, using N4-preprocessed images produced something closer, 𝒩 (0.56, 0.22), to the published values, 𝒩 (0.52, 0.18), reported in (54), resulting in a non-empty set for that cluster. This is consistent, though, with linear binning which does use N4 bias correction for preprocessing. We also mention that the Harvard Dataverse images used were preprocessed using N4 (3) which provides a third reason for its use on the University of Virginia image dataset (to maximize cross cohort consistency). In the case of the former image set, we did use the previously reported linear binning mean and standard deviation algorithm parameter values (i.e., 𝒩 (0.52, 0.18)). This was the only parameter difference between analyzing the two image sets.

**Figure 9:**
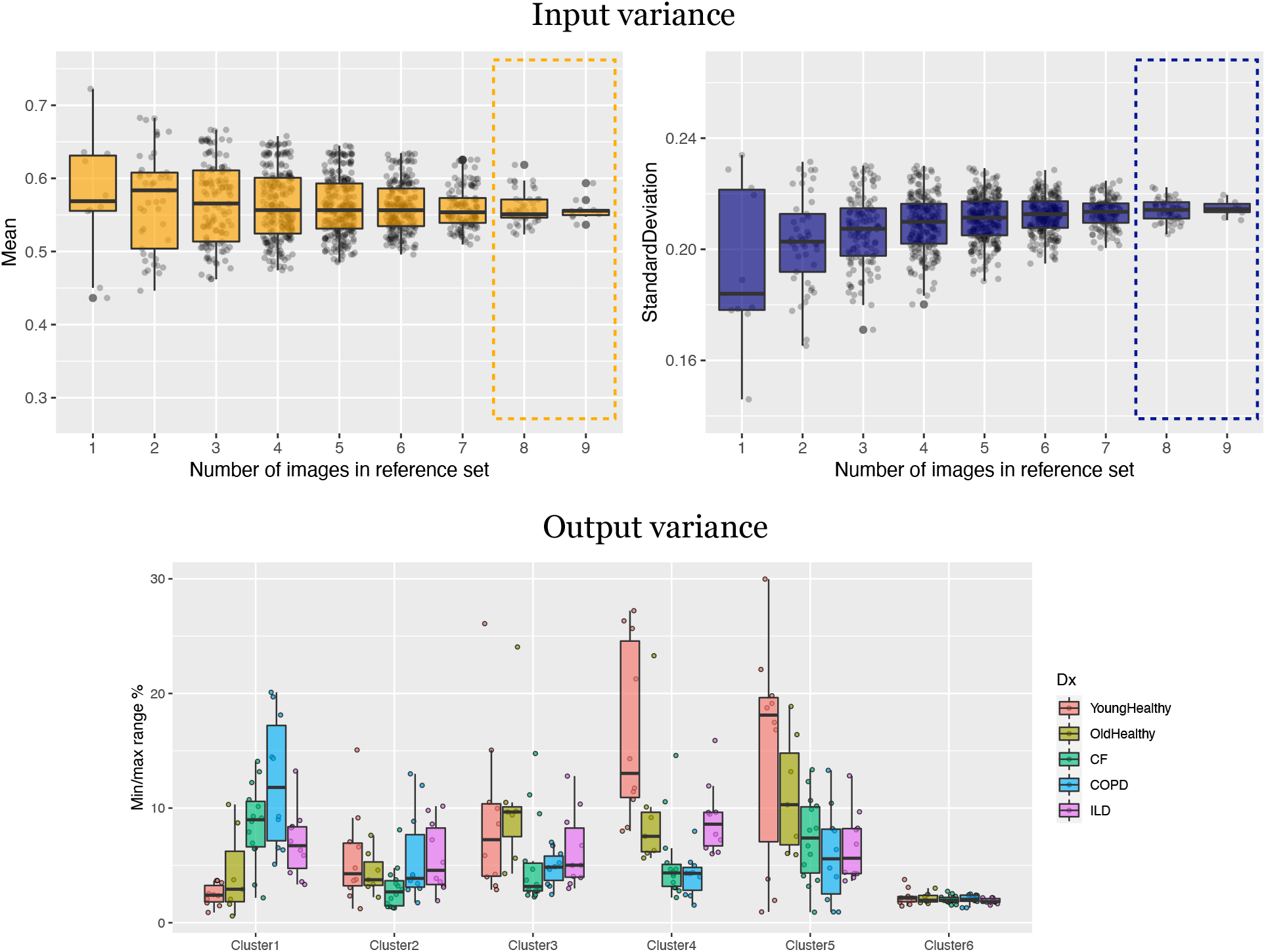
(Top) Variation of the mean (left) and standard deviation (right) over choice of reference set based on all different combinations of young healthy subjects per specified number of subjects. Although these parameters demonstrate convergence, there is still non-zero variation for any given set. (Bottom) This input variance is a source of output variance in the cluster volume plotted as the maximum range per subject as a percentage of total lung volume. We limit this exploration to reference sets with eight or nine images.

The previous implications of the chosen image reference set also caused us to look at this choice as a potential source of both input and output variance in the measurements utilized and produced by linear binning. Regarding the former, we took all possible combinations of our young healthy control subject images and looked at the resulting mean and standard deviation values. As expected, there is significant variation for both mean and standard deviation values (see top portion of Figure 9) which are used to derive the cluster threshold values. This directly impacts output measurements such as ventilation defect percentage. For the reference sets comprising eight or nine images, we compute the corresponding linear binning segmentation and estimate the volumetric percentage for each cluster. Then, for each subject, we computed the min/max range for these values and plotted those results cluster-wise on the bottom of Figure 9. This demonstrates that the additional requirement of a reference distribution is a source of potentially significant measurement variation for the linear binning algorithm.

### 3.4 Effects of MR-based simulated image distortions

As we mentioned in the Introduction, noise and nonlinear intensity artefacts common to MRI can have a significant distortion effect on the image with even greater effects seen with respect to change in the structure of the corresponding histogram. This final evaluation explores the effects of these artefacts on the algorithmic output on a voxelwise scale using the Dice metric (Equation (1)) which has a range of [0,1] where 1 signifies perfect agreement between the segmentations and 0 is no agreement.

Ten simulated images for each of the subjects of both the University of Virginia and Harvard Dataverse cohort were generated for each of the three categories of randomly generated artefacts: noise, nonlinearities, and combined noise and intensity nonlinearites. The original image as well as the simulated images were segmented using each of the five algorithms. Following our earlier protocol, we maintained the original Clusters 1 and 2 per algorithm and combined the remaining clusters into a single third cluster. This allowed us to compare between algorithms and maintain separate those clusters which are the most studied and reported in the literature. The Dice metric was used to quantify the amount of deviation, per cluster, between the segmentation produced by the original image and the corresponding simulated distorted image segmentation which are plotted in Figures 10 and 11 (left column). These results were then compared, on a per-cluster and per-artefact basis, using a one-way ANOVA followed by Tukey’s Honest Significant Difference (HSD) test. 95% confidence intervals are provided in the right column of Figures 10 and 11.

**Figure 10:**
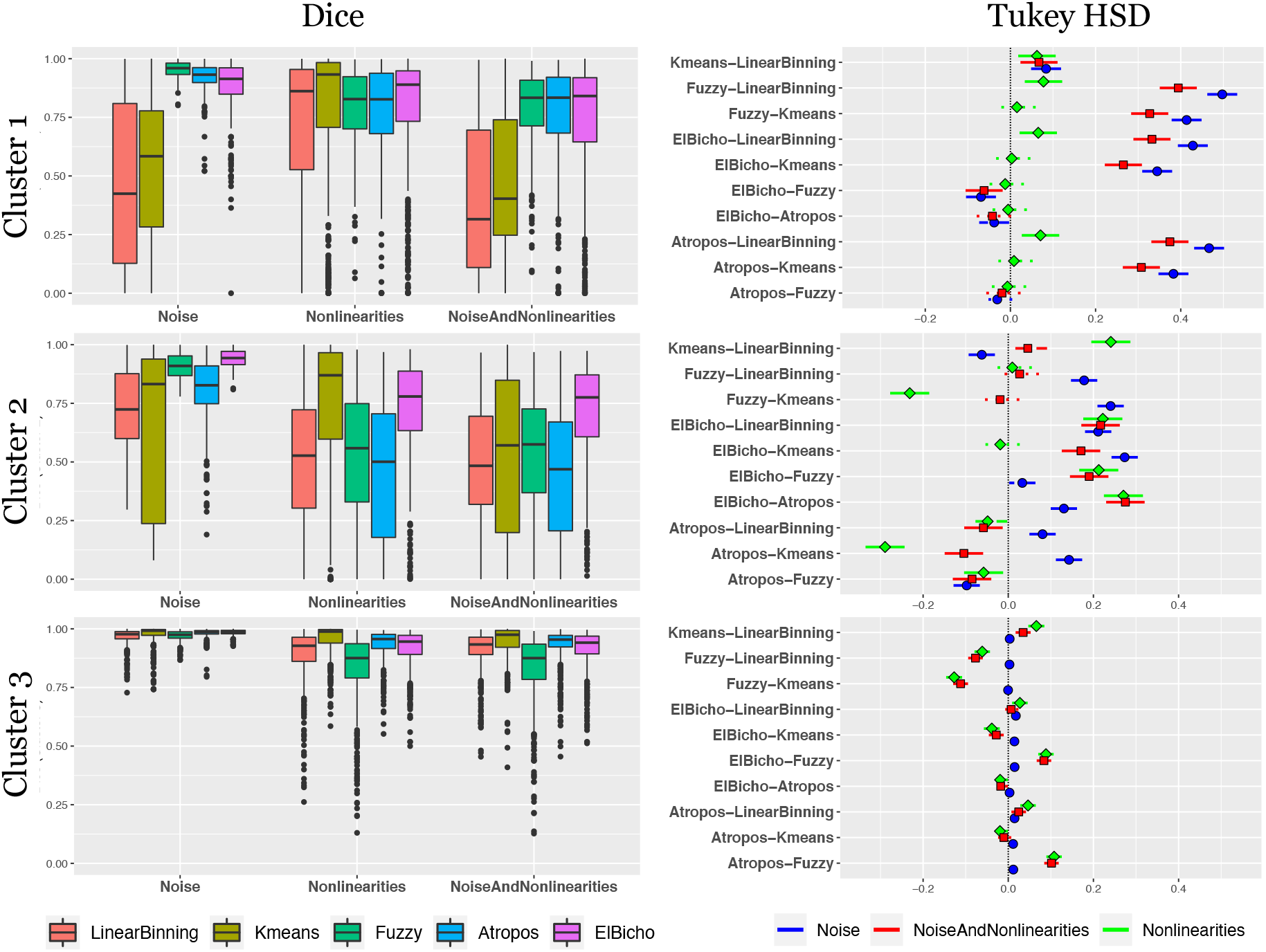
University of Virginia image cohort: (Left) The deviation in resulting segmentation caused by distortions produced noise, histogram-based intensity nonlinearities, and their combination as measured by the Dice metric. Each segmentation is reduced to three labels for comparison: “ventilation defect” (Cluster 1), “hypo-ventilation” (Cluster 2), “other ventilation” (Cluster 3). (Right) Results from the Tukey Test following one-way ANOVA to compare the deviations. Higher positive values are indicative of increased robustness to simulated image distortions.

**Figure 11:**
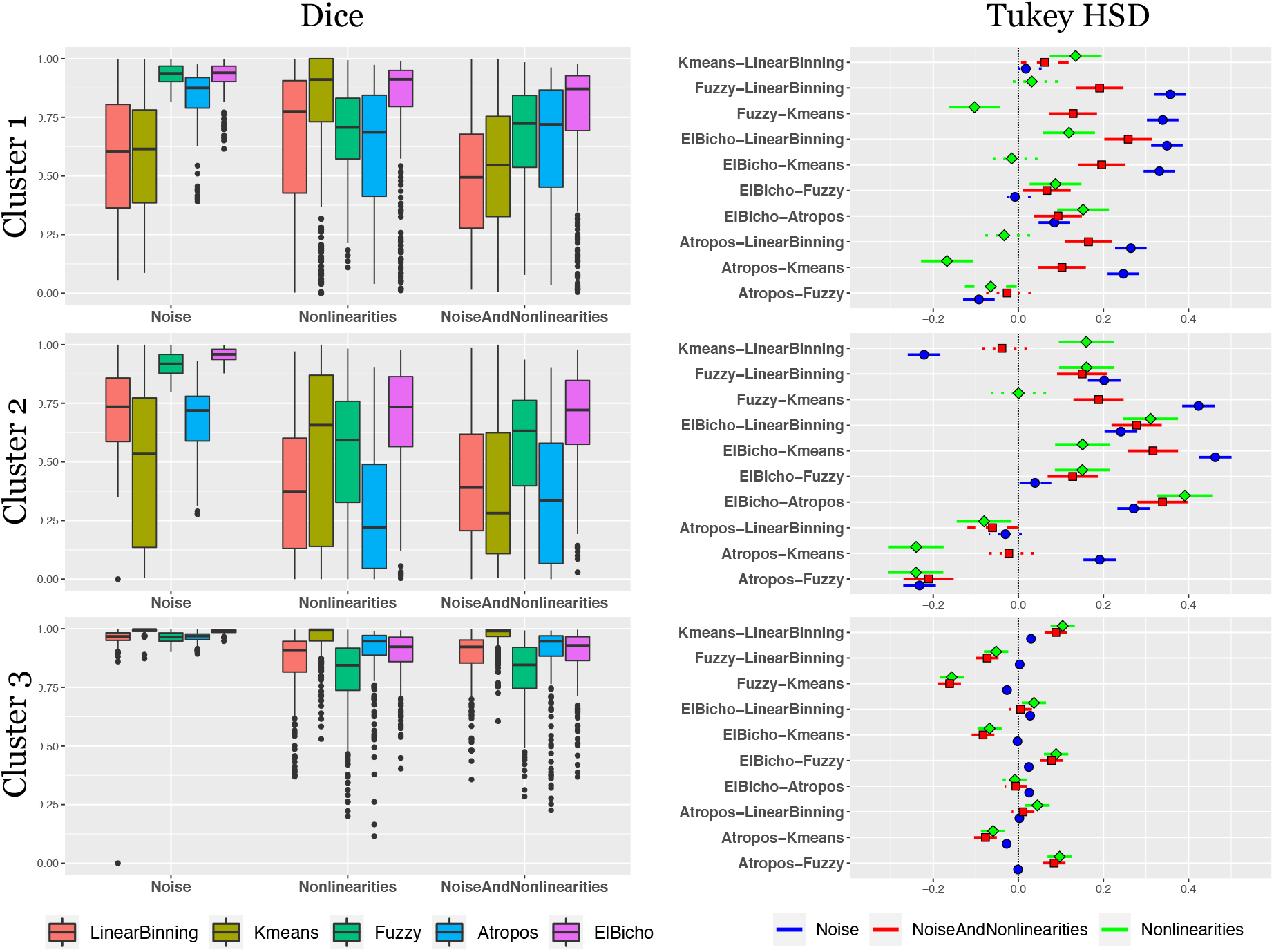
Harvard Dataverse image cohort: (Left) The deviation in resulting segmentation caused by distortions produced noise, histogram-based intensity nonlinearities, and their combination as measured by the Dice metric. Each segmentation is reduced to three labels for comparison: “ventilation defect” (Cluster 1), “hypo-ventilation” (Cluster 2), “other ventilation” (Cluster 3). (Right) Results from the Tukey Test following one-way ANOVA to compare the deviations. Higher positive values are indicative of increased robustness to simulated image distortions.

## 4 Discussion

Over the past decade, multiple segmentation algorithms have been proposed for hyperpolarized gas images which, as we have pointed out, are all highly dependent on the image intensity histogram for optimization. All these algorithms use the histogram information *primarily* (with many using it *exclusively*) for optimization much to the detriment of algorithmic robustness and segmentation quality. This is due to the simple observation that these approaches discard a vital piece of information essential for image interpretation, i.e., the spatial relationships between voxel intensities. A brief summary of criticisms related to current algorithms is as follows:

- In addition to completely discarding spatial information, linear binning is based on overly simplistic assumptions, especially given common MR artefacts. The additional requirement of a reference distribution, with its questionable assumption of Gaussianity and known distributional parameters for healthy controls, is also a potential source of output variance.
- Both hierarchical and adaptive k-means also ignore spatial information and, although they do use a principled optimization criterion, this criterion is not adequately tailored for hyperpolarized gas imaging and is susceptible to various levels of noise.
- Similar to k-means, spatial fuzzy c-means is optimized to minimize the within-class intensity variance but does incorporate spatial considerations which softens the hard threshold values and demonstrates improved robustness to noise. However, it is susceptible to variations caused by MR nonlinear intensity variation, similar to the GMM-MRF technique.
- The GMM-MRF approach does employ spatial considerations in the form of Markov random fields but these are highly simplistic, based on prior modeling of local voxel neighborhoods which do not capture the complexity of ventilation defects/heterogeneity appearance in the images. Although the simplistic assumptions provide some robustness to noise, the highly variable histogram structure in the presence of MR nonlinearities can cause significant variation in the resulting GMM fitting.

While simplifying the underlying complexity of the segmentation problem, all of these algorithms are deficient in leveraging the general modelling principle of incorporating as much available prior information to any solution method. In fact, this is a fundamental implication of the “No Free Lunch Theorem” (23)—algorithmic performance hinges on available prior information.

As illustrated in Figure 2, measures based on the human visual system seem to quantify what is understood intuitively that image domain information is much more robust than histogram domain information in the presence of image transformations, such as distortions. This appears to also be supported in our simulation experiments illustrated in Figure 10 and 11 where the histogram-based algorithms, overall, performed worse than El Bicho. As a CNN, El Bicho optimizes the governing network weights over image features as opposed to strictly relative intensities. This work should motivate additional exploration focusing on issues related to algorithmic bias on a voxelwise scale which would require going beyond simple globally based assessment measures (such as the diagnostic prediction evaluation detailed above using global volume proportions). This would enable investigating differentiating spatial patterns within the images as evidence of disease and/or growth and correlations with non-imaging data using sophisticated voxel-scale statistical techniques (e.g., similarity-driven multivariate linear reconstruction (1, 9)).

It should be noted that El Bicho was developed in parallel with the writing of this manuscript merely to showcase the incredible potential that deep learning can have in the field of hyperpolarized gas imaging (as well as to update our earlier work (56)). We certainly recognize and expect that alternative deep learning strategies (e.g., hyperparameter choice, training data selection, data augmentation, etc.) would provide comparable and even superior performance to what was presented with El Bicho. However, that is precisely our motivation for presenting this work—deep learning, generally, presents a much better alternative than histogram approaches as network training directly takes place in the image (i.e., spatial) domain and not in a transformed space where key information has been discarded.

Just as important, deep learning provides other avenues for research exploration and development. For example, given the relatively lower resolution of the acquisition image, exploration of the effects of deep learning-based super-resolution might prove worthy of application-specific investigation (10) (see, for example, ANTsRNet::mriSuperResolution). Also, with the same network software libraries, high-performing classification networks can be constructed and trained which might yield novel insights regarding image-based characterization of disease. One additional modification that we did not explore in this work, but is extremely important, is the confound caused by multi-site data which has yet to be explored in-depth. With neural networks, such confounds can be handled as part of the training process or as an explicit network modification. Either would be important to consider for future work.

## Data Availability

The University of Virginia imaging data used for the evaluation is available upon request and through a data sharing agreement. In addition to the citation providing the online location of the original He 2019 Dataverse data, a header-modified version of these data is available in the GitHub repository associated with this manuscript. Additional evaluation plots are also available at this location.

https://github.com/ntustison/Histograms

## Acknowledgments

Support for the research reported in this work includes funding from the National Heart, Lung, and Blood Institute of the National Institutes of Health (R01HL133889).

## Footnotes

1 The prior knowledge for histogram mapping is the general machine learning heuristic of clustering samples based on the minimizing within-class distance while simultaneously maximizing the between-class distance. In the case of k-means, this “distance” is the intensity variance.

2 A software codename designating a work-in-progress simply based on a shared admiration between the first and last authors of Portuguese futebol.

3 https://github.com/ntustison/Histograms

4 For completeness, we did run the same experiments detailed below using the uncorrected UVa images (and the previously reported parameters for linear binning) and the results were similar. These results can be found in the GitHub repository associated with this work.

5 This assessment is based on multiple conversations between the first author (as the co-developer of N4 and Atropos) and co-author Dr. Altes.

6 It is important to note that, although these experiments provide supporting evidence, our principal contentions stand prior to these results and are based on the self-evidentiary observations mentioned in the Introduction.

